# Evaluation of patient experience for a computationally-guided intranasal spray protocol to augment therapeutic penetration

**DOI:** 10.1101/2021.08.31.21262495

**Authors:** Saikat Basu, Uzzam Ahmed Khawaja, Syed A A Rizvi, BoLiang Gong, Waiman Yeung, Marcos A Sanchez-Gonzalez, Gustavo Ferrer

## Abstract

The global respiratory outbreak in the form of COVID-19 has underlined the necessity to devise more effective and reproducible intranasal drug delivery modalities, that would also be user-friendly for adoption compliance. In this study, we have collected evaluation feedback from a cohort of 13 healthy volunteers, who assessed two different nasal spray administration techniques, namely the vertical placement protocol (or, VP), wherein the nozzle is held vertically upright at a shallow insertion depth of 0.5 cm inside the nasal vestibule; and the shallow angle protocol (or, SA), wherein the spray axis is angled at 45° to the vertical, with a vestibular insertion depth of 1.5 cm. The SA protocol is derived from published findings on alternate spray orientations that have been shown to enhance targeted delivery at posterior infection sites, e.g., the ostiomeatal complex and the nasopharynx. All study participants reported that the SA protocol offered a more gentle and soothing delivery experience, with less impact pressure. Additionally, 60% participants opined that the VP technique caused painful irritation. We also tracked the drug transport processes for the two spray techniques in a computed tomography-based nasal reconstruction; the SA protocol marked a distinct improvement in therapeutic penetration when compared to the VP protocol.

## Introduction

For nasal inflammatory conditions, e.g. chronic rhinosinusitis (CRS), the single most-important delivery site for sprayed topical medication is the ostiomeatal complex (or, OMC)^1^ as it is the mucociliary drainage pathway and the dominant airflow exchange corridor between the main nasal cavity and the sinus appendages. For viral infections, e.g. SARS-CoV-2, the corresponding pharmaceutic target site during the initial infection phase is the nasopharynx^2-5^, with its tissue-level propensity of angiotensin-converting enzyme 2 (ACE2), a surface receptor that the virus binds to for cell intrusion. Evidence from *in silico* tracking in digitized medical scan-based geometries and *in vitro* measurements in 3D-printed anatomic replicas has confirmed^1^ that altering nasal spray protocols, e.g. by reorienting the nozzle axis, can often enhance drug delivery by multiple folds, especially for the posterior target sites, like OMC and the nasopharynx. To address the urgency induced by the COVID-19 pandemic for effective yet reproducible intranasal administration techniques, in this study we have tested patient experience for a representative new spray placement technique.

## Methods

Our study cohort comprises 13 healthy volunteers, recruited under an Institutional Review Board (IRB) approval. The subjects consented to assessing two different nasal spray placement techniques: (a) “vertical placement” protocol (or, VP), wherein the nozzle is held vertically upright at a shallow insertion depth of 0.5 cm inside the nasal vestibule; (b) “shallow angle” protocol (or, SA), wherein the spray axis is angled at 45° to vertical, with a vestibular insertion depth of 1.5 cm. The SA protocol represents a derivative of the “line-of-sight” (or, LoS) protocol recommended in published findings^1^ for CRS management. Figure 1(a)-(c) visually depicts the VP and SA protocols. The instructions were illustratively communicated (e.g., via Figure 1(a)-(b)) to the participants, and their feedback was recorded on a sensory attributes’ questionnaire. See Table 1 for the data.

**Table 1:**
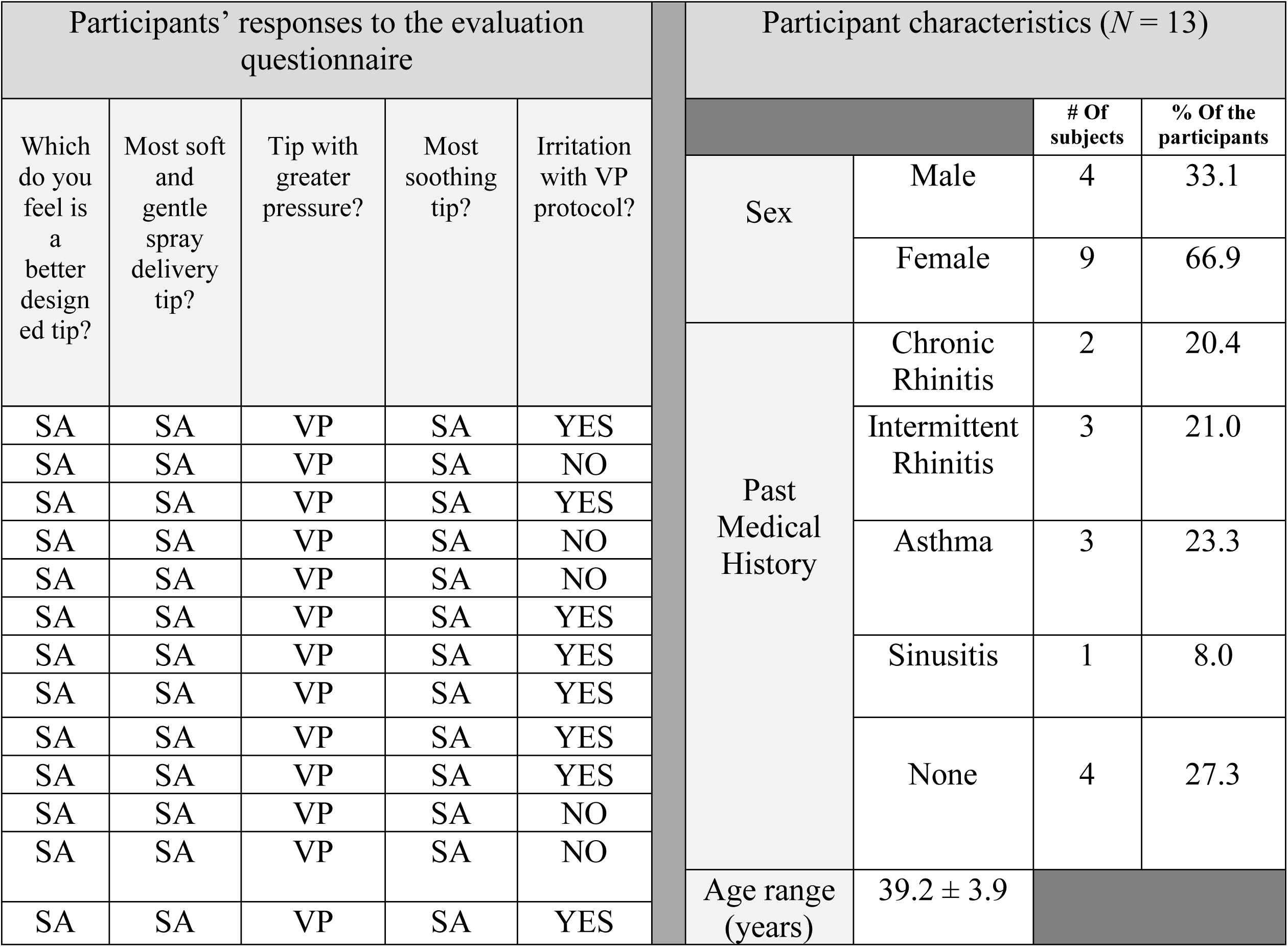
Nasal spray evaluation feedback from healthy volunteers

**Figure 1.**
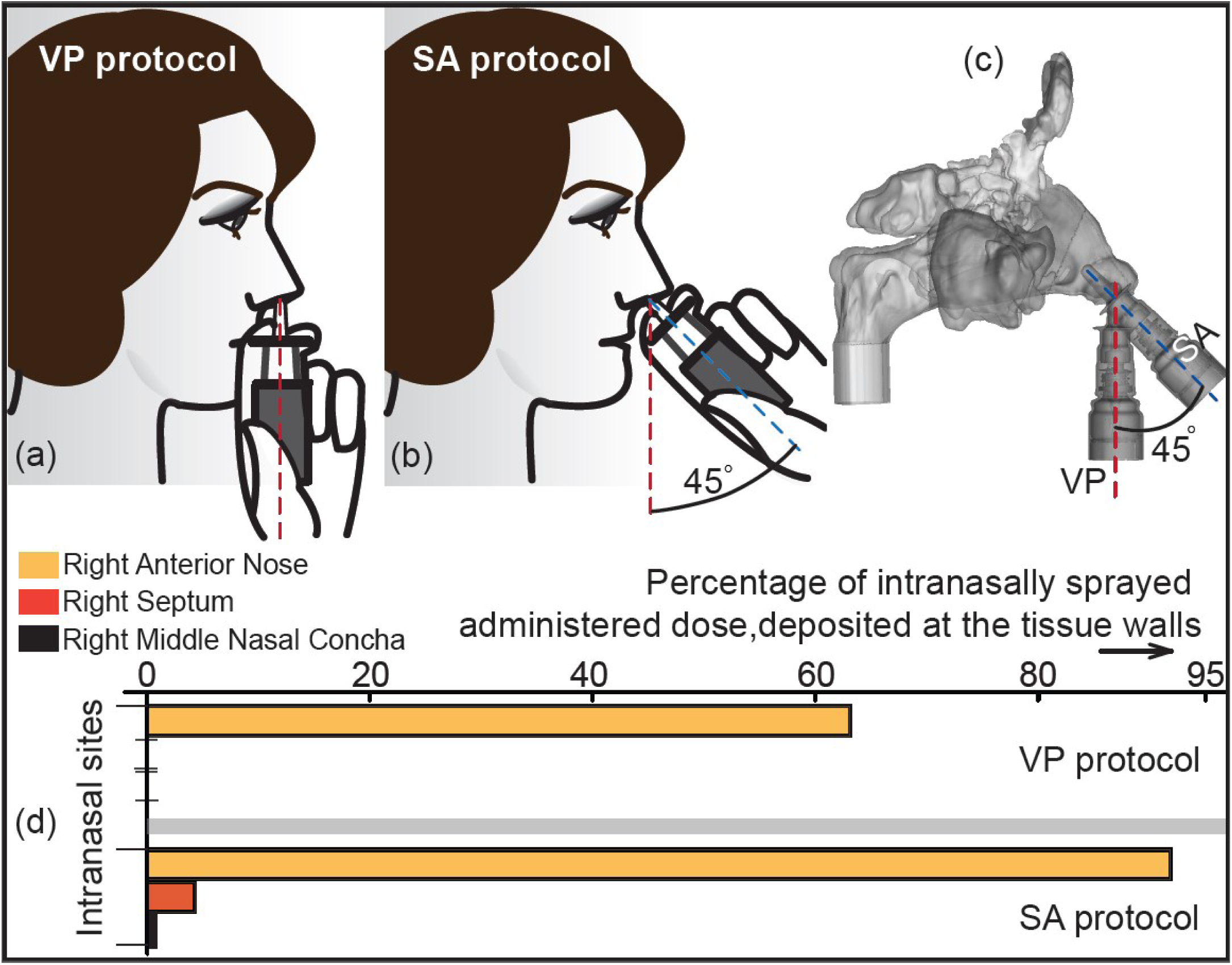
Comparison between VP and SA nasal spray protocols: Panel (a) shows the commonly used “vertical placement” (or, VP) protocol for nasal sprays. Panel (b) depicts the novel “shallow angle” (or, SA) protocol. Panel (c) presents a computed tomography based digitized geometry of the sinonasal airspace, with the VP and SA placements shown therein with a realistic spray bottle. Panel (d) compares the penetration for the VP and SA protocols in a representative anatomic geometry, comprising the right side of the *in silico* domain shown in (c). The VP technique reported a significant pharmaceutically-ineffective outflow through the nostril. The spray bottle axis orientations in the VP protocol and in the SA protocol are respectively marked by the red and blue dashed lines.

Experimentally-validated^1^ Computational Fluid Dynamics (CFD) simulations also helped check differences in sprayed delivery trends between VP and SA, by replicating inhalation in a computed tomography-based digitized airway reconstruction; see Figure 1(c) for the representative geometry. Retrospective *in silico* computational use of existing anonymized medical-grade imaging was approved with exempt status by the UNC Chapel Hill IRB. Scanned subject was a Caucasian female in her 20’s with BMI 32.6 and presented a clinical condition of CRS. We simulated normal steady breathing with inhalation rate of 22.30 L/min; the deviation from the measured rate (for the subject, via LifeShirts vests^6^) was < 0.2%. Details of the numerical scheme have been published separately^1^. Sprayed droplets were tracked against the ambient inspiratory airflow through discrete particle method with the droplet sizes following Rossin-Rammler distribution. Per *in vitro* measurements, the simulations implemented a half-cone angle of 31.65° and the droplet exit speed at nozzle was 10 m/s.

## Results

Table 1 details the volunteer evaluations for VP and SA protocols. All study participants reported that the SA protocol offered a more gentle and soothing delivery experience, with less impact pressure compared to VP. Furthermore, according to over 60% participants, the VP technique caused painful irritation. Consensus on the SA protocol was that it intranasally provided a comfortable mist-like sensation. Additionally, the CFD-based trends (see Figure 1(d)) confirm a distinct improvement in therapeutic penetration with the SA protocol.

## Discussion

While published *in silico* findings have established that targeted drug delivery to the posterior intranasal sites can improve significantly by perturbing the spray nozzle’s orientation and insertion depth; to our knowledge, our study is the first to collate *in vivo* data for a novel usage technique. Participant-reported unequivocally favorable experience with the SA protocol clearly justifies a full-scale clinical study to test medication compliance and therapeutic effectiveness for corresponding clinical conditions including allergic processes or viral illnesses with such spray parameters.

## Data Availability

The simulation data for the intranasal drug delivery trends in CT-based anatomic domains is available on-request from the corresponding author (SB).

## Acknowledgments

Basu was partially supported by the National Science Foundation, through the RAPID Grant 2028069. Any opinions, findings, and conclusions or recommendations presented here are, however, those of the authors and do not necessarily reflect NSF’s views.

## Author Contributions

*Concept and design:* Ferrer, Sanchez-Gonzalez

*Acquisition, analysis, or interpretation of data:* All authors

*Drafting of the manuscript:* Basu, Khawaja

*Critical revision of the manuscript for important intellectual content:* All authors

*Statistical analysis:* Ferrer, Sanchez-Gonzalez, Basu

*Administrative, technical, or material support:* Ferrer, Sanchez-Gonzalez, Basu

